# Influenza epidemic model with dynamic social networks of agents with individual behaviour: A self organize perspective

**DOI:** 10.1101/2020.08.11.20172114

**Authors:** L. López, M. Femández, L. Giovanini

## Abstract

It’s well known the existence of an interplay between the spread of an infectious disease like influenza and behavioural changes of individuals. An outbreak can trigger behavioural responses, at group and individual levels, which in turn can influence the course of the epidemic. Daily life interactions can be modelled by adaptive temporal networks in an explicit contact space through an agent based model, where each agent represents the interacting individuals. In this paper we introduce an individual based model where the behaviour of each individual is determined both by the external stimuli and its own appreciation of the environment and can be built as a combination of three interacting blocks: *i)* individual behaviour, *ii)* social behaviour and *iii)* epidemic state or epidemiological behaviour. We fit the model for a real influenza epidemic and perform the model validation, comparing the results with the classical approaches.

## 1. Introduction

Traditional epidemiological models represent the dynamics of infectious diseases through a non-spatial and population-based approach, where time has always been represented explicitly. These models do not explicitly address the causal factors for the development of epidemics. Instead, the size of the affected population is directly estimated. The development of an epidemic is represented as the change in the size of the segment of the infected population due to variations in the sizes of the other segments. These changes are supposed to be continuous and the rate of change at a given time is expressed by partial differential equations (Anderson et al., 1992).

This approach and its descendants take the population as the basis of modeling and provide a basis for modern epidemiology and a tool to assess the dynamics of epidemics and the health of the population. The models based on equations have a simple structure and a reduced number of parameters that allow an easy modeling process. By adjusting the parameters, the models can reasonably approximate the observed health data. For more than half a century, these traditional models have remained a support for epidemiology. In the last decade, however, these non-spatial models have attracted more and more criticism. Despite the advantages of these to predict health dynamics at the population level, they do not produce realistic and useful results for complex problems (Holmes, 1997; Cordero et al., 2004).

The models based on cellular automata arise to correct many of the flaws that classic models have. A cellular automaton is a dynamic system in which both time and space are modeled discretely. They are made up of a set of basic units called cells that are identical to each other. These models try to increase the heterogeneity of the population in order to produce more realistic results than traditional epidemiological models. One of the main limitations of them is that the units remain immobile in the grid, thus not contemplating the natural movement of the individuals of a population.

Agent-based modeling is a methodology that attempts to overcome the limitations of cellular automata. It uses virtual entities called agents as the basis of modeling. Traditionally, agents have been used in the resolution of problems in a distributed way, where each agent has the ability to solve a part of the problem but you can only reach the global resolution by working together. More recently, agents have been used to perform simulations, allowing modeling of individuals with complex interactions and behaviors within a dynamic environment. There are some works that goes in this direction.

Epstein et al. (2002) simulates an *SEIR* epidemic assuming a constant contagion rate in two possible infectious states (one low and one high infection rate), two cities of 400 each are modeled. Each city contains 100 houses with two working adults and two children in each. In each city there is a school and a workplace. All the children go to school in their city. Most adults go to work in their city, while a small fraction works in the neighboring city. There is only one hospital used by both cities, with 5 workers from each city.

On the other hand Teweldemedhin et al. (2004) implements a model in which all elements of the system, including the environment, are implemented as agents. A controlling agent initiates the simulation and supervises the creation or deletion of the other agents, as well as their evolution. A statistical agent collects all the information obtained from the simulation and performs calculations with it. Agents that simulate individuals move through a continuous space and interact with others by choosing one or more partners at random from their neighbors.

Yergens et al. (2006) also implements all the elements as agents, but in this case, individuals, cities and roads that unite them are modeled in a flexible and detailed manner. It is exemplified for the case of bird flu, but the system can be adapted to other cases. A representation based on graphs is obtained, with the nodes representing the cities and the arcs with associated weights representing the roads. Individuals interact with each other differently before and after becoming infected. This part of the hypothesis of a change of habits of the individual to know that he is sick. Individuals can move from one city to another and disperse the disease, also in adjustable form

Commonly, the necessary conditions for infections to persist in host populations have been defined through thresholds. The maintenance of infectious agents depends as much on the demographic rates as on the size of the population. In systems where population density or seasonal migrations cause the abundance of hosts and the relevant demographic rates fluctuate (Lloyd-Smith et al., 2005) and also the risk of disease is influenced by individual factors (age, sex), rumps (territoriality, behavior of group) this poses particular challenges. Spatial effects such as the diffusion of individuals (Davis et al., 2008) or the metapopulation structure (Lambert et al., 2013) may be important for the persistence of the infectious agent.

Of the current models, methods based on reproductive number have been used to explore the maintenance in multiple hosts in human-animal interfaces (Radivojac et al., 2013) and identify the specific contributions of each species to the transmission (Nishiura et al., 2009; Streicker et al., 2013). These dynamics that are not in equilibrium can be crucial for persistence, especially in systems with temporarily variant outbreaks.

In this paper we introduce a new model where the individual is the basic modelling unit and the overall dynamics of the system arises from the aggregation of individuals dynamics. The behaviour of individuals is a key to understanding the epidemiological system, it determines how the unit unfolds in the environment and therefore defines its overall dynamics. We propose to model the behaviour of individuals in a modular fashion such that their conduct is defined as the aggregation of three parts: *(i)* individual behaviour, *(ii)* social behaviour and *(iii)* epidemiological behaviour.

The work is organized as follows: Section 2 make a general description of the model, introducing the conceptual framework; Section 3 describes the way in which the model was implemented; Section 4 shows the model’s fitting and parameter’s estimation; Section 5 show and analyze the results obtained, validating the model in both numerical and visual way and finally, Section 6 summarize the conclusions of this work and propose future worlines.

## 2. Model formulation

Complex systems have many components that interact in a non-linear way, these systems can not be represented and analyzed with traditional modeling methods. One way to explain and know its characteristics is through spatially explicit modeling based on individuals. The approach has four characteristics that make it an attractive alternative when modeling an epidemic: *(i)* Agents are autonomous, that is to say that there are no external mechanisms that control the behavior of the same, it is governed by internal rules. *(ii)* They have social capacity, that is, agents are able to interact with each other. *(iii)* They are reactive, that is to say that they react to external stimuli coming from the environment or from other agents and act according to said stimuli.

An infectious disease outbreak can trigger behavioural responses, at group and individual levels, which in turn can influence the course of the epidemic. To formulate models in which infectious disease dynamics and social behaviour are interindepent, we need to incorporate the mechanism behind any mutual influence, answering the next questions: To what extend do people themselves, their social group, media opinion or personal perceptions influence individual behaviour? and, how are the perceptions that determine behaviour influenced by properties of an infection? Following the idea proposed by Gross and Blasius (2008), we propose a model based on adaptive co-evolutionary network framework where the interplay between the dynamic of the epidemic process and the temporal evolution of the structure of the network, that model the social interactions, is considered. An individual-based model is built on agents whose behaviour emerge from the interactions of three intertwined components: *i)* Individual behaviour, *ii)* social behaviour and *iii)* epidemic state.

The idea behind this decomposition of the agent behaviour is to facilitate the description of the system and to elucidate the interrelationship between individual behaviour and disease spreading. Besides, the modular structure employed to model the agent’s behaviour allows to use different tools for each component.

i. The social behaviour establishes how an individual relates with others. It is given by intraspecific relationships like communication and social practices. It is determined by the individual behaviour but it is also modified by the environment where the individual lies.
ii. Epidemiological structure is how the inherent factors of the causative agent interact, whether physical, chemical or biological, environment and guest in a population in a given time period. The epidemiological characteristics are the result of the epidemiological structure and are expressed by the frequency and distribution of disease in the population. According to the epidemiological characteristics, the epidemiological structure is dynamic, modified according to the behaviour of the disease in the community. The epidemic state determines individual’s health evolution. The epidemiological analysis considers the structure, epidemiological characteristics and behaviour of a disease. To model this aspect we can choose to incorporate epidemiological stages and deterministic or stochastic functions that determine the transition between them
iii. The individual behaviour determines how an individual react to the factors that influence their behaviour like communication, cultural norms, personal circumstances and alternatives, among others. It is given by a combination of emotions, perceptions and experiences of the agent. Emotions play an important role in affecting memory, attention and reasoning. Human intelligence can not only be rational thinking and logical reasoning, but also has strong emotional capacities. Psychologically speaking, emotions represent the reaction of an individual for an evolution of their own internal state and the interactions between them and their environment (Damasio, 1994)

As its seen there is a feedback between social and individual behaviour. The individual interact with other members of the group and modifies its perceptions and experiences, which make him react in a given way that modifies the group dynamic. The presence of a disease modifies the behaviour of each individual, which modifies the structures of the group (Figure 1). Social structures evolution depends on individual’s dynamics. A feedback loop is created between group dynamics (local interactions) and social (global structures).

**Figure 1:**
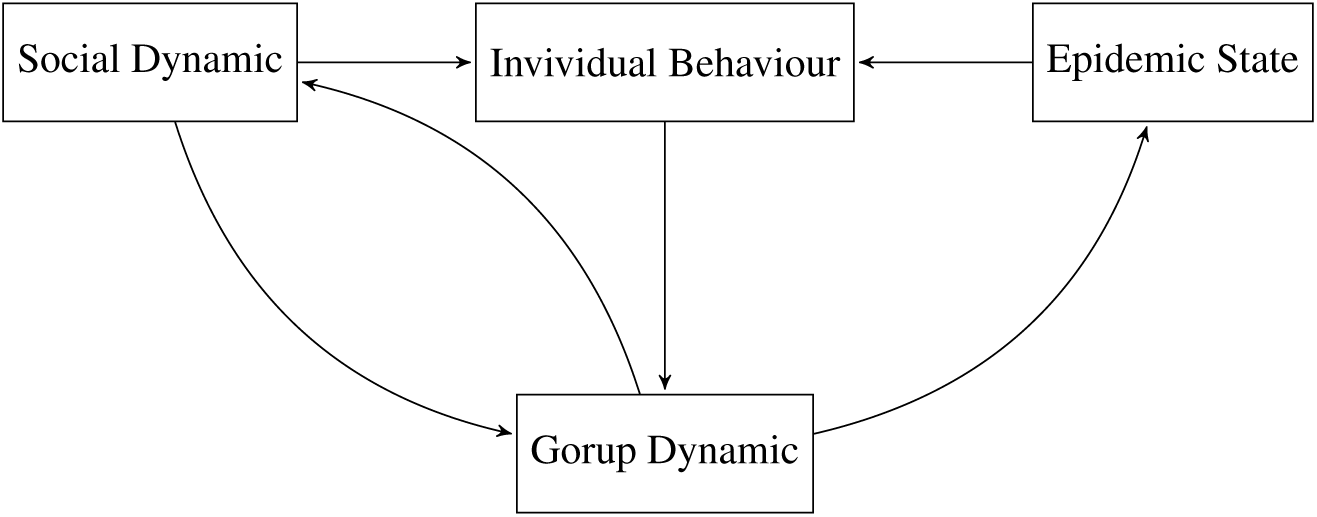
Adaptive network scheme.

### 2.1. Individual behaviour

Human behaviour can be influenced by countless factors ranging from media and person-to-person communication to personal feelings and perceptions. The behavioural towards an infectious disease is determined by a combination of these influences and people evaluate them responding their alternatives.

Human emotions psychologically can be classified into three levels: *(i)* primary emotions or those considered as an individual intrinsic responses to external stimuli, *(ii)* secondary emotions, those activated when the primary emotions are connected with the current and past perception and *(iii)* senior emotions those produced by the course of long-term social contacts in a given environment (Damasio, 1994; Xiao-Juan and Wei-Ren, 2007).

Fuzzy Cognitive Maps (FCM) provides a powerful tool based o graph theory to model behaviour system in terms of interacting concepts representing a state or a characteristic, through linkages that express their relationship in a hierarchical structure. A FCM integrates experience and accumulated knowledge of the system as a result of the method by which it is generated, ie using human experts who know how the system works under study with all its components under different circumstances based on statistical measures. Concepts are nodes and interconnections between concepts and represents causal relations between them. These maps model the dynamics of a complex system as a collection of concepts and cause-effect relationships between these concepts (Figure 2).

**Figure 2:**
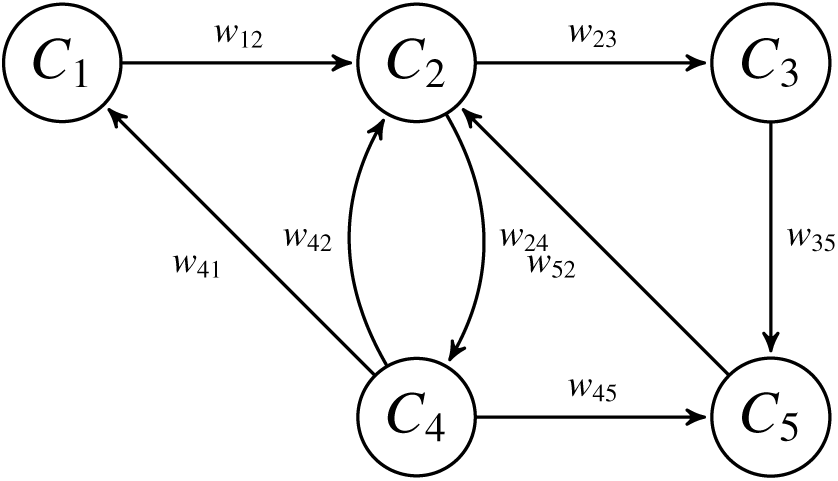
Fuzzy Cognitive Map diagram

Interconnections between different concepts are characterized by a weight *w_ij_* which describes the degree of causality between two concepts. Weights can take values in the interval [−1,1]. The sign of the weight indicates the type of causality: *w_ij_* > 0 indicates positive causality between *C_i_* and *C_j_* while *w_ij_ <* 0 indicates negative causality between *C_i_* and *C_j_*. Positive causality means that when *C_i_* increase *C_j_* will also increase. On the other hand, negative causality means that when *C_i_* increase *C_j_* will decrease. When there is no relationship between two concepts, then *w_ij_ =* 0. The value of *w_ij_* indicates the degree of influence between the concepts *C_i_* and *C_j_*. Generally, the value of each item is estimated by calculating the influence of other concepts on the specific concept (Lee et al., 2012; Mei et al., 2014).

### 2.2. Social behaviour

Social behaviour determines how an individual relates with other individuals. It is related to individual behaviour establishing a feedback relationship. In a multi-agent system, a set of individuals (agents) develop and interact in an environment (Iantovics, 2010). Simulations with multi-agent systems allow direct simulation of individuals, their behaviors and their interactions, as well as other elements of the system (Epstein, 2006). There are properties at the individual and system level. The properties at individual level correspond to the behaviors and characteristics of each agent. System-level properties are the global properties of the environment in which agents live, and emerge as a result of the action and interaction of agents with each other and with the environment (Epstein, 2006). From the dynamic point of view, multi-agent systems can be analyzed as a set of *N* mobile agents with a simple dynamic given by:

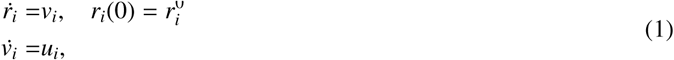

where *r_i_* = (*x_i_, y_i_*)*^T^* is the position in space, 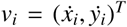 is the velocity and 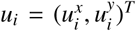 is the control entry thats determine the behaviour of the agent *i*. The environment can be modeled as agents whose behavior is predefined, for example immobile agents in the case of obstacles. In addition, this way of modeling behavior allows us to incorporate the effects of perception and communication between agents. The individual behavior of each agent in relation to the others (agents and environment) and their objectives is given by *w_i_*, assuming that each agent interacts only with those agents that are within a range of interaction *r* (Figure 3). Then, the behavior of the systems arises from the aggregation of the individual behaviors of the agents that compose it (self-organization). The idea of self-organization rests on the premise of *individual rationality* (Lee, 2011; Cristiani et al., 2015), a doctrine in which each individual pursues the best possible outcome for himself. In this way, each individual tries to do the best for himself without taking into account the effects of his actions on the rest of the group. In this sense, the behavior of the system results from the superposition of the individual behaviors, emergent behavior of the system, which is more complex than the sum of the individual behavior (Helbing et al., 2011).

**Figure 3:**
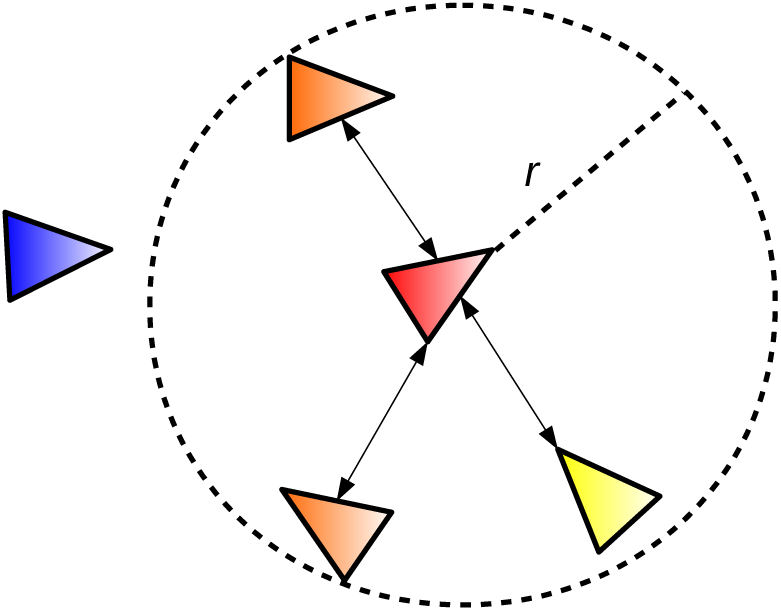
Agent’s neighborhood

Emergent behavior differentiates between micro and macro levels in a self-organized process. It is considered that of the local interactions between the components of a network (micro level) (Figure 3) emerges a structure or global pattern (macro level).

### 2.3. Epidemiological state

The epidemiological state determines the health state of each individual with respect to epidemic states. They can be modelled using a finite state automata that includes all the information available about the disease behaviour. This way of modelling allows to mix stochastic and deterministic transitions between states, improving the modelling capabilities. Furthermore, it allows modelling different epidemiological situations (quarantine, vaccination, multiple strains, among others) by modifying the automata (changing the states and their transitions) and/or including individual heterogeneity (modifying the parameters of the automata), while it retains the simplicity.

Formally, each stochastic finite state automata in the network can be defined as 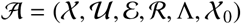, where 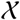 is a finite states set, 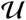 is the input set, 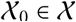 is the initial state, 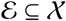 is the output set, 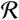 are the rules or transition function between states. and Λ is the output function. The transition function 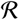 indicates which state is going to happen knowing the current state and the symbol being read. The transition rules can be deterministic or stochastic. These rules can be represented as a transition matrix. The rate or probability of going to a certain state to another may depend on the current state and the entry. The output value may also depend on an own distribution function probability of each state or entry. Transitions by empty entries are also possible in this case, it may be associated probability state change. When this automaton simulate the following value must be calculated randomly, following a probability distribution (Mikler et al., 2005).

## 3. Model Implementation

In section 2 we introduce the model’s general structure, describing the different blocks that characterize individual’s behaviour. In the present section detail about the model implementation is given. Here we provide an explanation on which tools were used to model the different aspects of the agent’s behaviour. As we previously introduce in the previous section these modules are:

- **Epidemic state** is implemented through a stochastic Moore machine where each state is one of the possible epidemiological states (susceptible, exposed, infected, among others);
- **Social behaviour** is modelled by a individual based model implemented as an Agent Based Model;
- **Individual behaviour** is modelled through a *FCM* that emulates the emotional system of individuals.

### 3.1. Epidemic state

The epidemic state is defined as a Moore machine given by 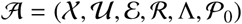, where 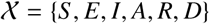 is the finite set of states, comprising six epidemic states: *S* (susceptible), *E* (exposed), *I* (infectious symptomatic), *A* (infectious asymptomatic), *R* (recovered) and *D* (dead or empty). 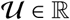 is the input set.

An individual only receives an active input when it is in state *S*, emitted by another in state *I* or *A*, when the receiver is connected to the sender in the network. The state transition function is defined as 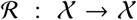, applied to the active state at iteration *t*, probabilistically decides the active state at iteration *t* + 1. The function is applied in two steps, one corresponding to the change of states by infection and recovery and another corresponding to the movement. If movement is not simulated, then the second step is not performed. In order to decide the state changes two probability matrices are defined, one for the transition on empty input (Table 1), where μ is the probability of natural death, γ is the probability of recovery, and another for the transition by contact with infectious (Table 2) in which taking into account the neighborhood size v and the input value as 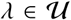. 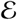 is the output set, the output is non-zero when the state of the node is *I* or *A*.

**Table 1:**
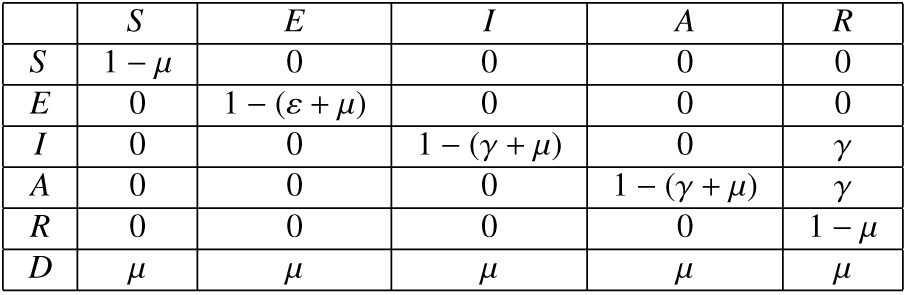
Transition matrix

| | $S$ | $E$ | $I$ | $A$ | $R$ |
| --- | --- | --- | --- | --- | --- |
| $S$ | $1 - \mu$ | 0 | 0 | 0 | 0 |
| $E$ | 0 | $1 - (\varepsilon + \mu)$ | 0 | 0 | 0 |
| $I$ | 0 | 0 | $1 - (\gamma + \mu)$ | 0 | $\gamma$ |
| $A$ | 0 | 0 | 0 | $1 - (\gamma + \mu)$ | $\gamma$ |
| $R$ | 0 | 0 | 0 | 0 | $1 - \mu$ |
| $D$ | $\mu$ | $\mu$ | $\mu$ | $\mu$ | $\mu$ |

**Table 2:** Transition matrix by contact.

| | $S$ | $E$ |
| --- | --- | --- |
| $S$ | $1 - \lambda/\nu$ | $\lambda/\nu$ |
| $E$ | 0 | 1 |

The transition matrix are derived from the classic model parameters (Chowell et al., 2006). These parameters are deterministic in the classical model, but as a result of an aggregation of individual probabilities under the hypothesis of a sufficiently large population. Therefore, it is the probabilistic transition that supports the classical model and not the other way around. If 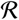 is applied directly to each node in the set 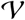, there is a decreasing variability as the population increases, until it coincides with the deterministic evolution of the classical model. The initial distribution of individuals in the environment can be random, fulfilling the hypothesis of homogeneous distribution for large population sizes. The model also allows exploring different spatial distributions for individuals as well as the number of individuals. Potentially infectious contact occurs between infectious individuals (symptomatic and asymptomatic) and susceptible individuals.

The output function 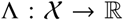 outputs the infection rate of the automata if it is in *I* (*β*) or *A (q β*, being *q* the probability of infection by contact with an asymptomatic individual) said rate is non-zero. This value is related to the *β* of the classic model, but it is not exactly the same, it is the probability of transmission by contact. The potential number of infectious contacts *c* is related to the size of the neighborhood which determines the connectivity with other nodes in the network, but it is not exactly the number of contacts. In order to perform the equivalence between the two methodologies, the number of contacts in the network is multiplied by the density of cells occupied in the grid (which may vary according to the grid size used and the number of individuals to be simulated). The combination of the *β* probability with the potential number of contacts *c* is what will result in the infection rate of each infectious individual. As the value obtained from the parameter setting, the *β* value is obtained from the choice of the neighborhood size and the *β* parameter used in the classical model, using the *β* = *βc*. In the Figure 4 the state transition graph of the automata that models the epidemic state is shown.

**Figure 4:**
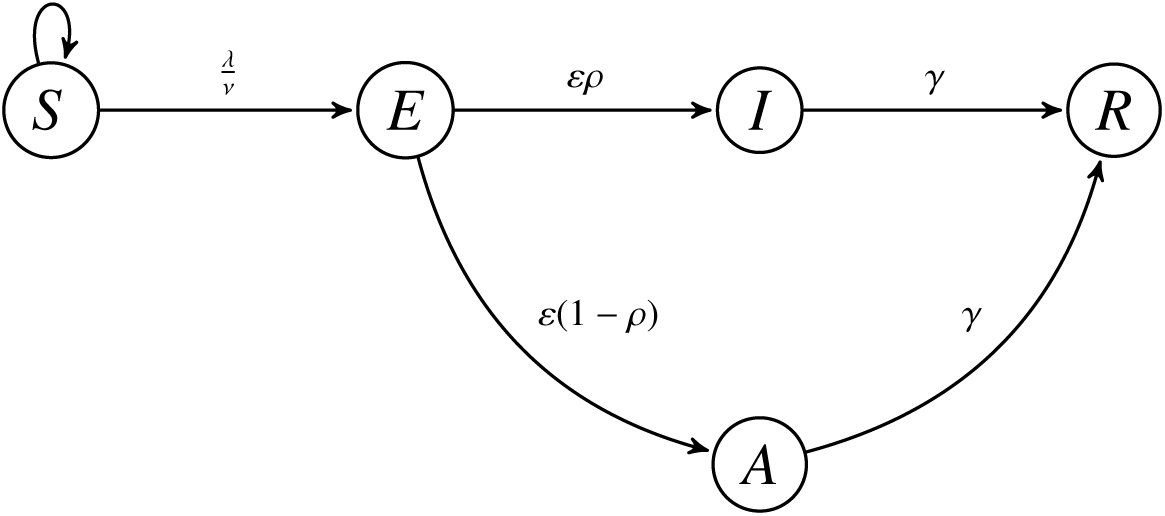
State graph of the epidemic states model

The initial state vector 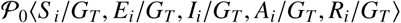 indicates the probabilities of each state being the initial state of the automaton. Defining as *G_T_* the total number of cells in the grid and *S_i_*, *E_i_*, *I_i_*, *A_i_*, *R_i_*, *D_i_* as the initial number of individuals in each state in the grid (Its sum being equal to *G_T_* and not to the total population, since *D_i_* includes the empty cells. In our case, we only consider probabilities for the states *S*, *E*, *I* and *D*, derived from the initial values of individuals in the classical model, and the cells that are left empty accordingly. All other states do not have initial individuals.

### 3.2. Social behaviour

Social behaviour establishes how an individual relates with other individuals having a main influence in the structure and dynamic of the social interactions. The social behavior of agents is modeled through self-organization, that is, defining the behavior of each agent, then the behavior of the system emerges from the aggregation of the behavior of all agents. A possible way to define the behavior from the rules introduced by Reynolds (Reynolds, 1987).

- keep neighboring agents as close as possible;
- avoid collisions of neighboring agents;

These rules are known as the rules of cohesion, separation and alignment. The main problem with the implementation of these rules is that they can be interpreted very loosely. The interpretation of these rules was described in later works by Reynolds (Reynolds, 1999, 2000). Considering a group of *N* agents whose dynamic behavior is given by

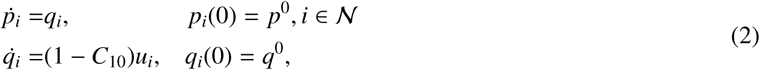

where *u_i_* is the entry that determines the behavior of each agent and is affected directly by the output of the *FCM* by the term (1 − *C*10), thus limiting the movement of the agents. Recent works have focused on Lagrangian concepts (Mogilner et al., 2003; Cucker and Smale, 2005; Olfati-Saber, 2006), which focuses on the relationship between the individual. The input *u_i_* is obtained from the gradient of a potential function that is automatically generated from the superposition of the individual agent potentials. Therefore, the actions of each agent is not only determined by local information, but is also a product of the combination of information available throughout the system. In this way the movement of an agent *i* is described by the equations (2). The mobile agents have the tendency to keep a distance *d* > 0 from their neighbors within a previously defined interaction radius, forming the structures defined as lattices or lattices. In a system without obstacles, each agent receives an entry that can be summarized as follows:

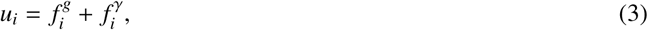

where 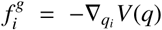 is the term that determines the cohesion of the group and 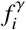 is a feedback function that determines the objective of the group.

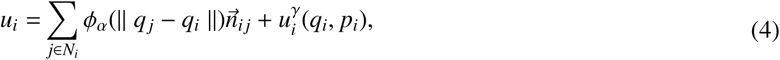

where *p_i_* is the velocity component of the agent and 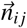 is a vector along the line joining *q_i_* and *q_j_* and is given by the following expression:

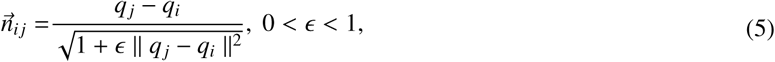

which defines the protocol of behavior between agents. The term 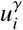 is the navigation function defined as:

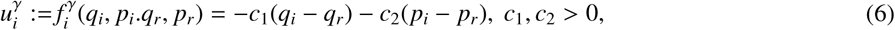

the pair (*q_r_*, *p_r_*) ∊ ℝ^2^ is the state of a static or dynamic agent type that is called *γ*-agent and represents a target of the group. If the *γ*-agent is a dynamic agent, its movement is defined by the system of equations similar to (2)

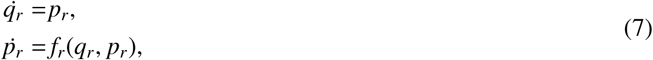

when the *γ*-agent is static, then it has a static state 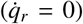 that is retained for every instant of time. The collective dynamics of a group of agents whose behavior is given by the rules established in (4) has the following form:

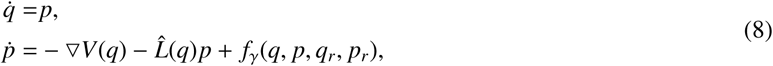

where *V*(*q*) is the collective potential defined in (14) and 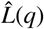 is the Laplacian of the graph *G(q)* with adjacency matrix dependent on the position *A*(*q*) = [*a_ij_*(*q*)] One of the characteristics of this type of systems is the tendency to form groups on the part of the individuals that compose it as a consequence of the dynamics that define the behavior of each one. Let *r* > 0 be the range of interaction between two agents that determines the set of spatial neighbors of an agent *i* and is defined as:

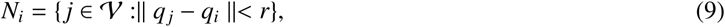

where ||.|| is the Euclidean norm in ℝ^2^. Given an interaction range *r* > 0, a spatially induced graph 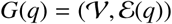 can be described by 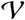 and the set of connections 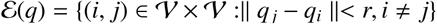, which clearly depends on *q*. The graph *G*(*q*) is defined as a network structure (*G*(*q*), *q*) is defined as a network framework. If the interaction range for all the agents in the network is the same, the *G*(*q*) network becomes an unmanaged network. The lattices are defined with the aim of designing protocols that allow group dynamics. This idea motivates the following algebraic restrictions between the agents:

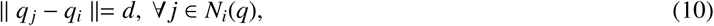

An *α–*lattice is a *q* configuration that satisfies the constraints on (10), where *d* is the scale and *k = r/d* and the radius of the *α*–lattice. It must be taken into account that the network induced by a α-lattice is not required to be connected. A quasi *α*–lattice is a *q* configuration that satisfies the constraints:

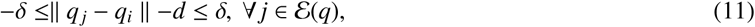

where *δ* ≪ *d* is the measure of connection uncertainty.

To measure to what degree a *q* configuration differs from a *α–*lattice, the following energy function is used:

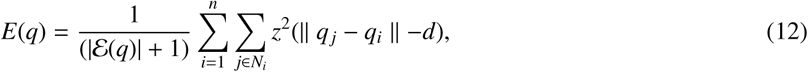

where *z*^2^ is the potential of pairs. You can use different functions as potentials according to the behavior you want to represent. This measure of energy can be seen as a potential for a *n* particle system. For quasi-*α–*lattices the energy of a *q* configuration can be calculated by:

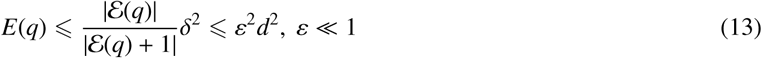

The potential function *V*(*q*) for a group of agents is a non-negative function *V* ℝ^2^*^n^* → ℝ_≥0_ with the property that any solution of the set of algebraic constraints in (10) is closely related to a local minimum of the function *V*(*q*) and vice versa. The way in which these two quantities are related is measured using the deviation energy that acts as a function of distance. A collective potential can be seen as a deflection energy function with an even scalar potential that has a finite cutoff. The potential function for a *q* configuration can be defined by:

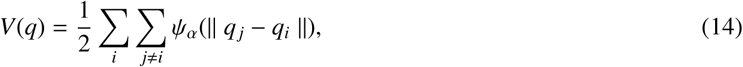

where *ψ_α_* defines the atraction-rejection potentialof the a-lattice it’s defined as a mapping rule *ψ*(*z*): ℝ_≥0_ → ℝ_≥0_ where *z* is the dimension.

### 3.3. Individual behaviour

The individual behaviour is modeled through a *FCM* based on the model proposed by Mei et al. (2014). In this model the concepts *C_i_* with *i* = 1,2,…, 10, are divided into three groups:

i. **Input concepts**, where *C*_1_ is near infected individuals, *C*_2_ is near recovered individuals and *C*_5_ is knowledge of the global epidemic situation, representing the perceptions that the agent receives from the environment (primary emotions);
ii. **Internal concepts** representing emotions and feelings of the individual, ie, secondary emotions, where *C*_3_ is the health state of individual (given by the epidemic behaviour), *C*_4_ is knowledge of local epidemiological situation, *C*_6_ is the assessment of local and global epidemiological situation, *C*_7_ is optimism level, *C*_8_ is the memory of similar situations and *C*_9_ instant reactions;
iii. **Output concept** corresponding to the senior emotions (*C*_10_) representing the actions taken by the agent as a result of a decision process.
iv. **Inputs** *u_i_*, where: *u*_1_, is the density of infected individuals that have contact with a given individual; *u*_2_ is the density of recovered individuals that have contact with a given individual; *u*_3_ is the epidemic state given by the epidemic behaviour and *u*_4_ is the knowledge of the local epidemiological situation.

Figure 5 shows the connections between the concepts *C_i_* and inputs *u_i_* with the associated weights. The value of *C*_10_ limits the number of contacts make by each individual within the environment, which affects the control entry *u_i_*(*t*). The values of the weights *w_ij_* of the matrix *W* are detailed in Table 3. The values of the input concepts *C*_1_ and *C*_2_ are estimated at each iteration as density of infected individuals Neighborhood size (*ν*) and density of recovered over neighborhood size respectively. The value of the concept *C*_3_ is given by entry *u*_3_ and is determined by the exit of the Moore machine that models the epidemic state of the individual

**Figure 5:**
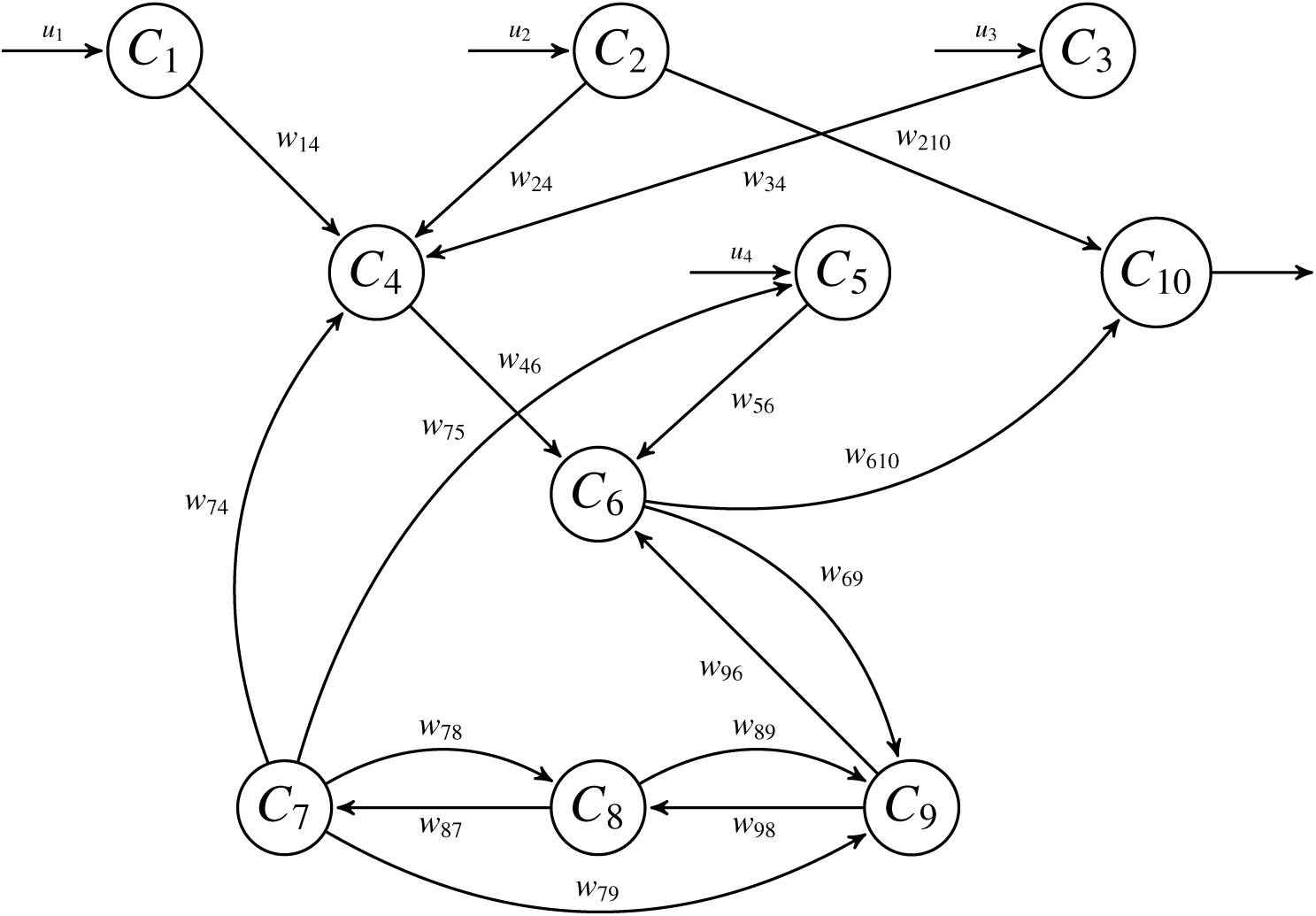
Fuzzy Cognitive Map used to model the individual behaviour for flu epidemics.

**Table 3:**
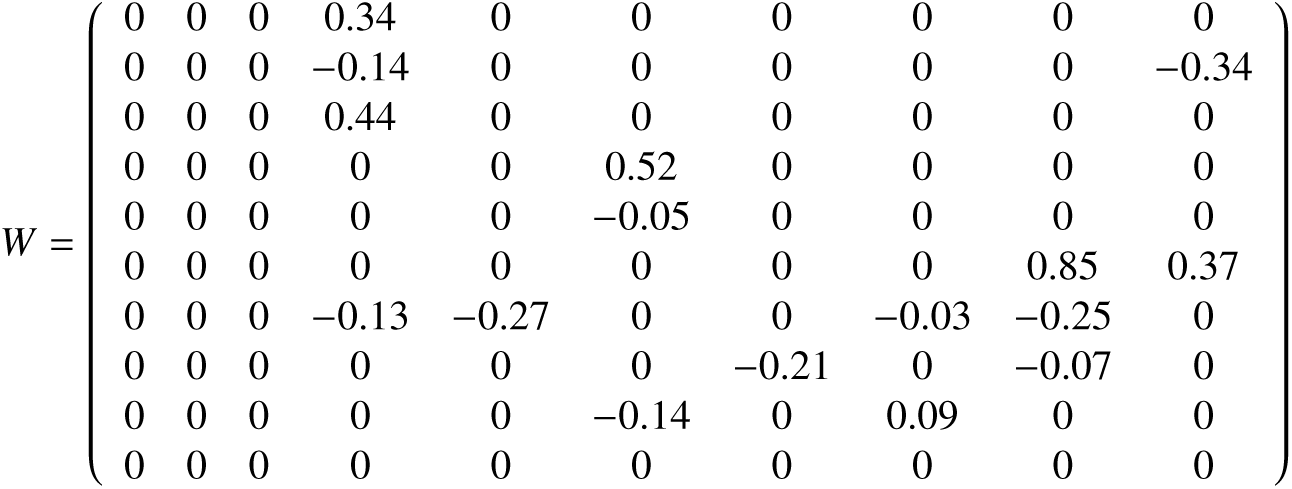
*FCM* weight matrix.

The value of *C*_10_ quantifies the individual’s perception of the overall epidemiological situation. In the work of Lee et al. (2012), authors use a function to calculate the probability of infection according to the value of *C*_10_. In the model implemented in this paper *C*_10_ affect the control input *u_i_*(*t*)as it’s shown in equation 2.

### 3.4. Framework implementation

Algorithm 1 summarize the in general terms the model implementation. Given the initial configuration of the contact network, the evolution of the system depends on the interaction of each of the blocks that model the behaviour. At first, the number of contacts of each individual is determined by the *FCM*, then a remap of his contact network through the extraction and replacement algorithms is made and then the epidemic state is updated.

In Algorithm 1 can be seen that for each individual in the environment each one of the behavioural blocks is performed. First of all the initialization of the whole system is made as shown in algorithm 2.

In the individual behaviour actualization step the *FCM* concepts are updated giving the input concepts *C*_1_, *C*_2_, *C*_3_ and *C*_5_. The new concept values are computed according to:

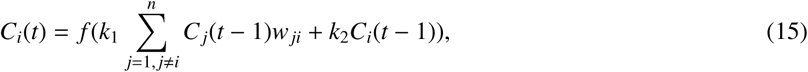

#### Algorithm 1

Model’s dynamic algorithm

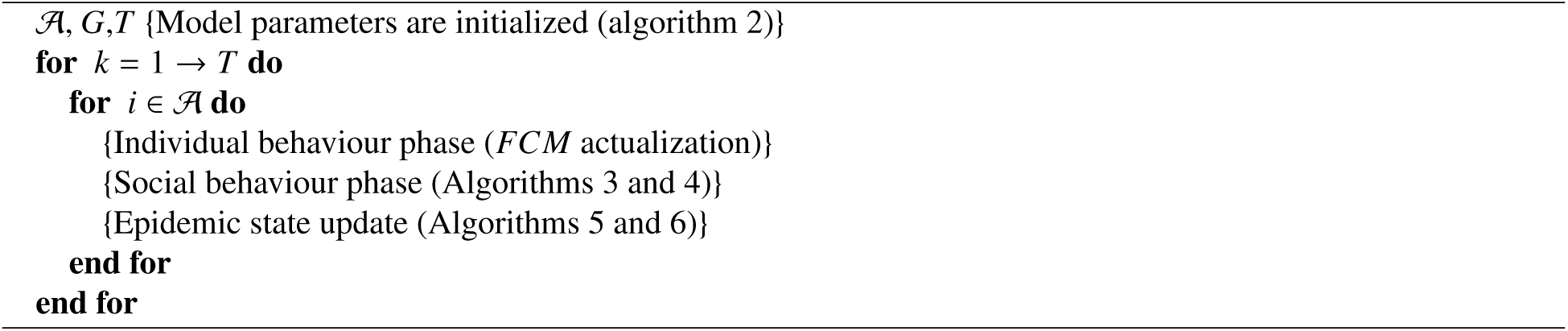

#### Algorithm 2

Set initial parameter

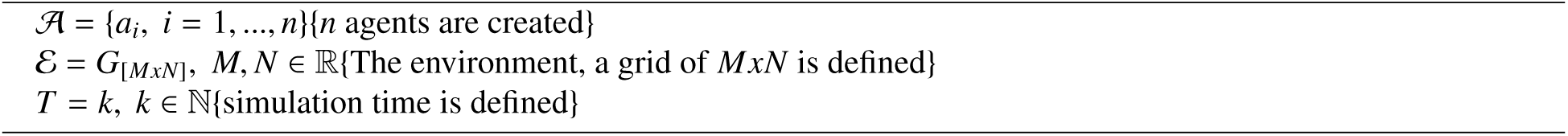

where *k*_2_ is the contribution of the previous value of the concept in the calculation of the new concept and *k*_1_ is the influence of the interconnected concepts in the configuration of the new value of the concept *x*_1_. The two parameters *k*_1_ and *k*_2_ satisfy 0 < *k*_1_, *k*_2_ < 1. The threshold function *f* used is:

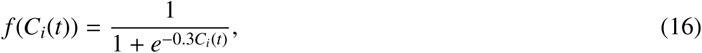

After the initialization phase comes the *Individual Behaviour Phase* in which the actualization of the *FCM* following

#### Algorithm 3

Infectious state

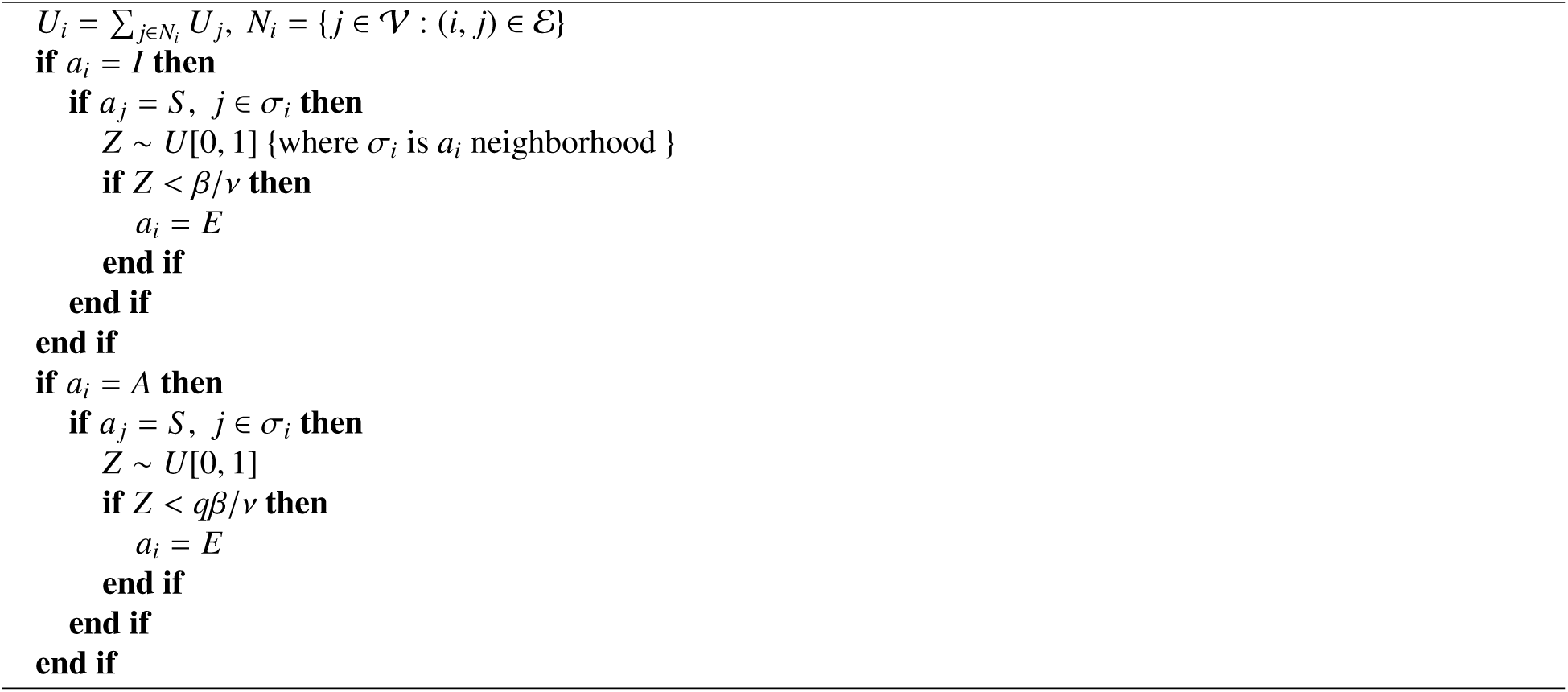

the algorithms previously detailed is made. Following this step, the next one is the *Social Behaviour Phase* is the next one, where algorithms 3 and 4 are performed.

Finally, the last phase is the *Epidemic state update* in which algorithms 5 and 6 are performed.

To summarize the process, in the figure 6 you can see a flow diagram in which they are explicitly stated which are the different blocks of behavior that are executed in each step of simulation. In this diagram it serves to have a better overview of how the different components of the proposed framework are related. Next we test the capacity of the model with a real case and proceed with the validation stage of it.

**Figure 6:**
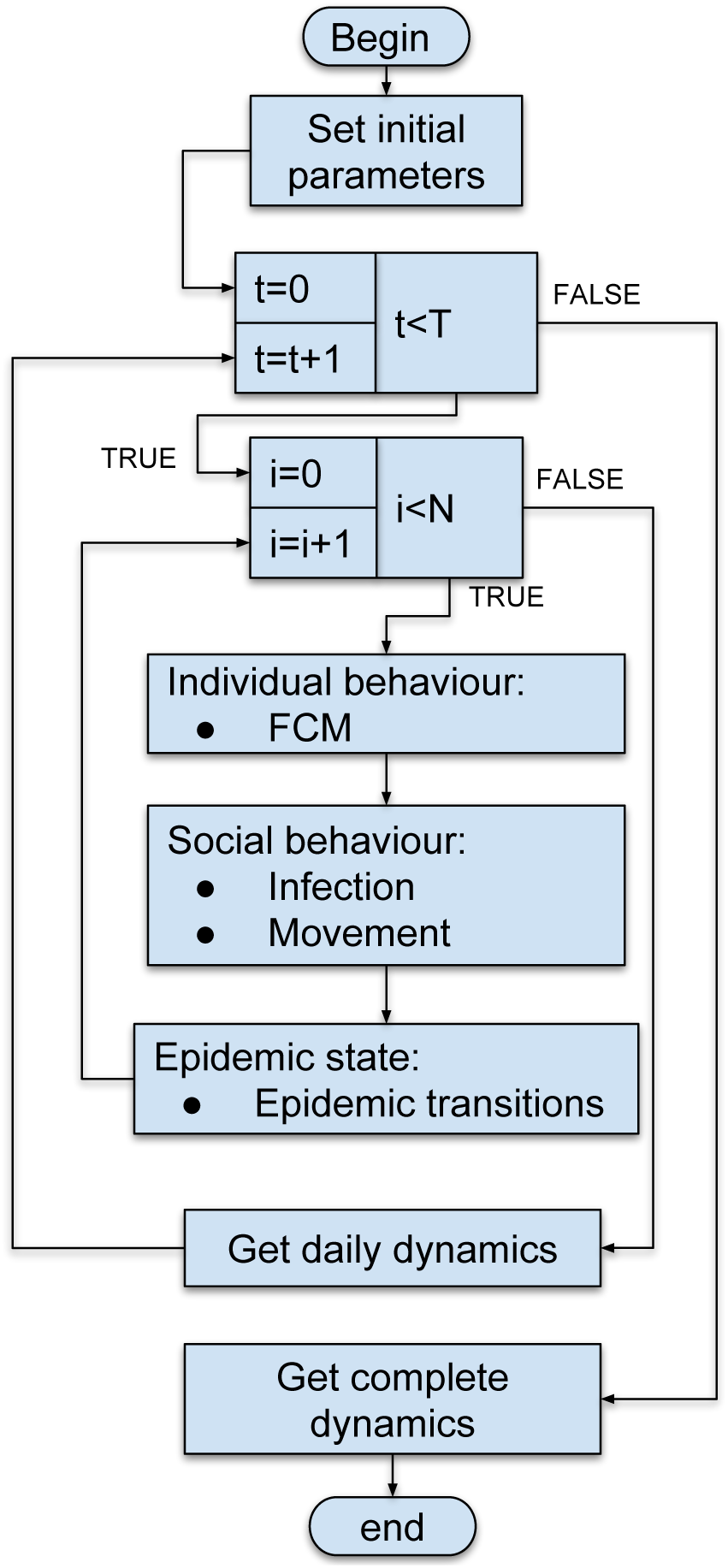
Framework’s flux diagram

#### Algorithm 4

Movement algorithm

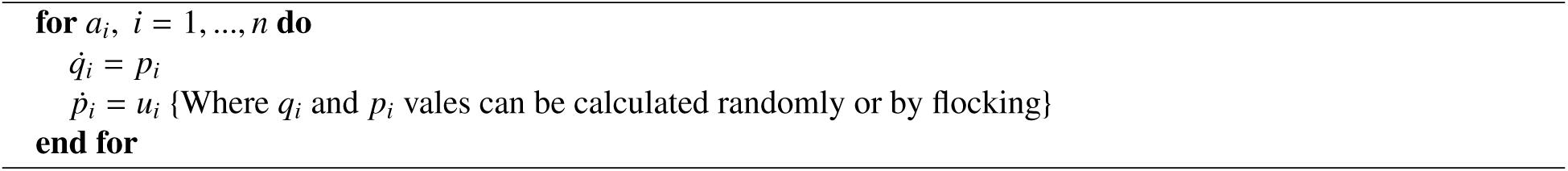

#### Algorithm 5

Exposed state

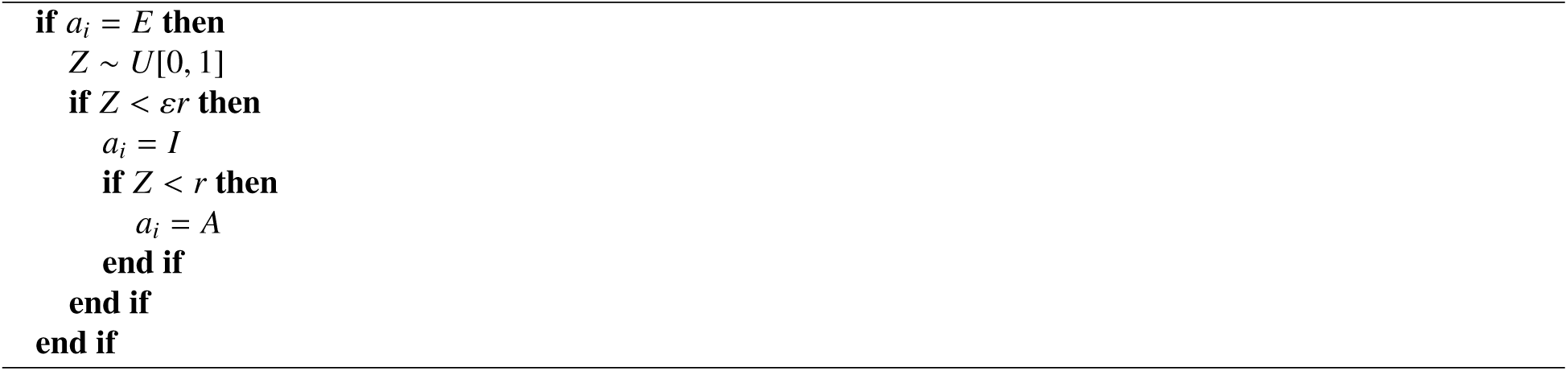

## 4. Model evaluation

To evaluate the model we analyze the Spanish flu in the Swiss canton of Geneva in 1918 (Chowell et al., 2006). We use this data set because this epidemic has been extensively studied and the availability of data. Due to the error function is not completely convex the model parameters were estimated following a two step procedure: *i)* using a global stochastic optimization method more precisely simulated annealing (Deb et al., 2002) and then *ii)* gradient based local optimization algorithms (Byrd et al., 2000). This procedure allows us to explore the entire parameter space looking for good candidates (global search), which are used to find the best parameters for the model through a local search. Stochastic optimization methods provide good starting points for optimization methods based on gradient. The objective function used was the normalized square error (NMSE)

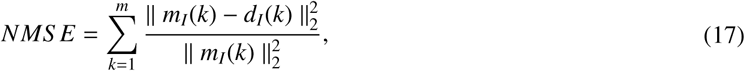

where *m_I_*(*k*) are the temporal dynamics obtained by the model (infected individuals) and *d_I_*(*k*) are the real dynamics.

Table 4 shows the fitted model parameters where *β* is the transmission rate, *ρ* is the proportion of infected reported infectious, *γ* is the recovery rate, *α* is the rate of diagnosed, *q* is the rate of infection of asymptomatic individuals, *N_e_* is the initial population of exposed individuals, *N_i_* is the initial population of infectious individuals and *r* is radius of the neighborhood used. Due to of the great difficulty of having time series describing influenza epidemics, the data used is from the 1918 Spanish flu registered in the Swiss city of Geneva has been widely studied (Chowell et al., 2006).

**Table 4:** Model parameters

| $\beta$ | $\rho$ | $\gamma$ | $\alpha$ | $q$ | $N_e$ | $N_i$ | $r$ |
| --- | --- | --- | --- | --- | --- | --- | --- |
| 8.1 | 0.085 | 0.246 | 0.465 | 0.3 | 207 | 136 | 3 |

### Algorithm 6

Recovery state

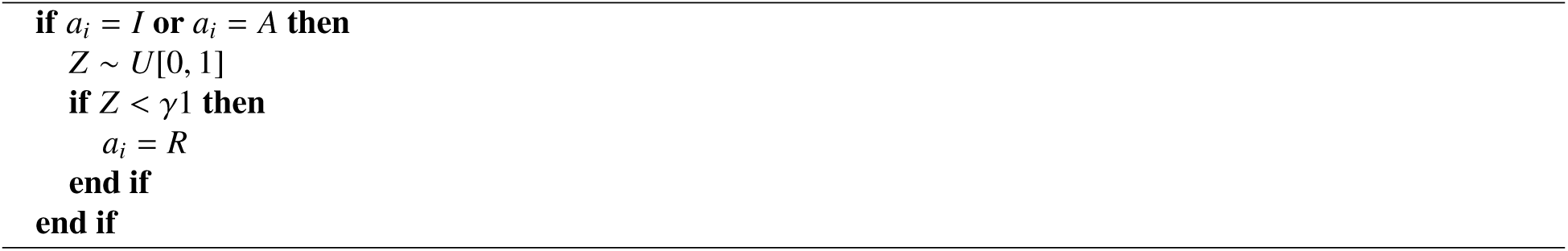

For FCM’s training we use the algorithm proposed by Mei et al. (2014). The value of each concept is calculated taking into account the influence of other concepts over the specific concept taking into account the value of causal relationships between them. As we are modelling a local outbreak (city level), the value of *C*_5_ = 0.5 which is the equivalent to a phase 4 alert according to World Health Organization. It is characterized by verified human-to-human transmission at community-level. Another aspect to consider is the threshold function to use in for the actualization of concepts *C_i_*. The sigmoid function is usually used with this purpose. One of the characteristics of the sigmoid function is that its domain is the interval (−∞, ∞), while its range is (0,1), ie it can not get 0 or 1 as a value for any concept. What is more, a sigmoid function with is not a good normalizer in [0,1], because the slope of the function in *x* = 0 is very different from the slope in *x* = 1 depending on the value of λ so this function is not the most suitable for use as a normalizer (Lee et al., 2012). For this reason we chose to use a linear function as a threshold function. A linear function would not be appropriate for the inference process of an FCM because it is monotonically increasing in the interval (−∞, ∞). However, if we know the range of the domain during the inference process, it can become an excellent option since it considers the ends of the inference interval and retains the same slope in that interval (Lee et al., 2012). For this reason a function of the form is proposed,

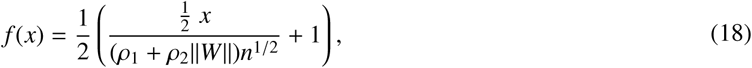

where *ρ*_1_ and *ρ*_2_ are values in [0, 1], *W* is the weight matrix and *n* is the number of concepts. The matrix *W* obtained is

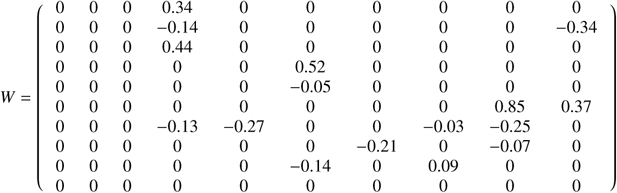

In order to validate the obtained matrix 10000 vectors of concepts 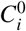 were generated, where the value of each element *C_i_*, *i* = 1,…, 10 was generated randomly within the interval [0,1]. The results show that the values of the concept *C*_10_ final are always within the range (0,1). When you try to reproduce the classical model with the minimized parameters, various obstacles and use alternatives are presented. First, the problem of scale use is presented. It is evident that the larger the grid and employed population, the closer we hypothesize the classical model large population (and the greater the computational cost of the simulation). As the extension of the validity of this hypothesis is just something that is being tested by applying the model of automata, is neither necessary nor desirable to use grid sizes too large when analyzing the temporal behaviour of the epidemic, but it is for the purpose of validating the model using the classical model parameters.

## 5. Results

Model validation is one of the most important steps in the process of construction. You can use two methods to carry out this process. On one hand, graphical methods illustrate a wide range of complex aspects of the relationship between the model and observed data. Furthermore, numerical methods accurately illustrate particular aspects of the relationship between the model and observed data. Numerical methods for validation of a model show a single number or result representing the same degree of reliability. Figure 7 shows the model fit against the classic SEIR model. Both models capture the real dynamics very well as shown in the figure approaches its very similar to the actual data. The error calculated for the classic SEIR model, according to *NMSE* function is 3.3, while for the proposed model is 1.6.

**Figure 7:**
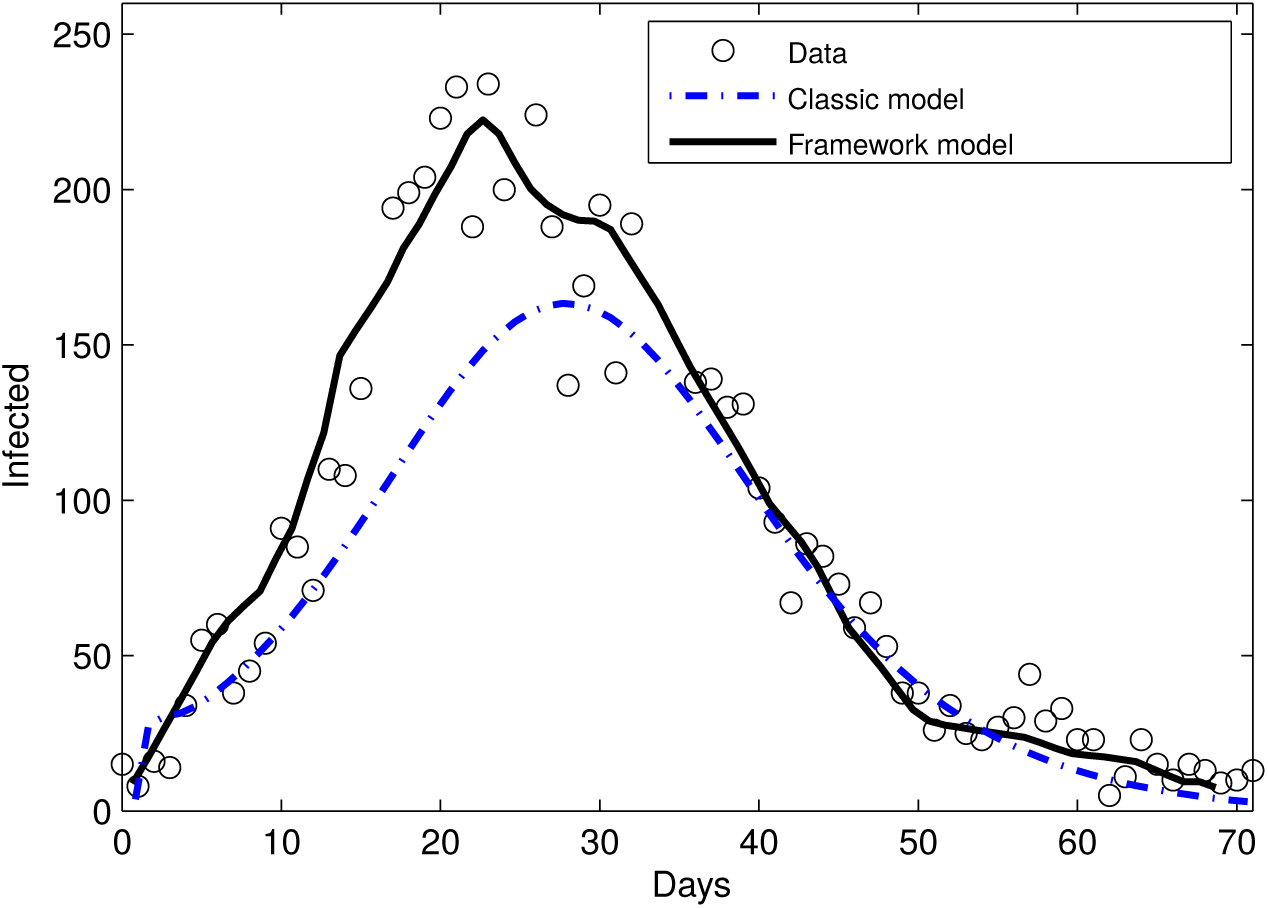
Model fitting. Mean over 500 simulations.

### 5.1. Visual validation

In order to verify the ability of our model to adjust to real epidemiological dynamics, the results of the adjustment are shown in Figure 7, there can be observed the adjustment of the spatially explicit model based on mobile individuals with and without social behavior against the adjustment of the classic *SEIR* model. For both cases it is observed that there are some points corresponding to the real data that do not fit precisely and it can be observed that both the model with social behavior and the model without social behavior capture the peak of the epidemic wave accurately while the classic model no. As a model represents a stochastic approach we made 1000 experiments in order to capture the average behavior.

Figure 8a shows the residuals of the models and allows to evaluate the quality of the same to approximate the real dynamics. On the other hand Figure 8b shows the distribution of the errors committed by each model including the classic model.

**Figure 8:**
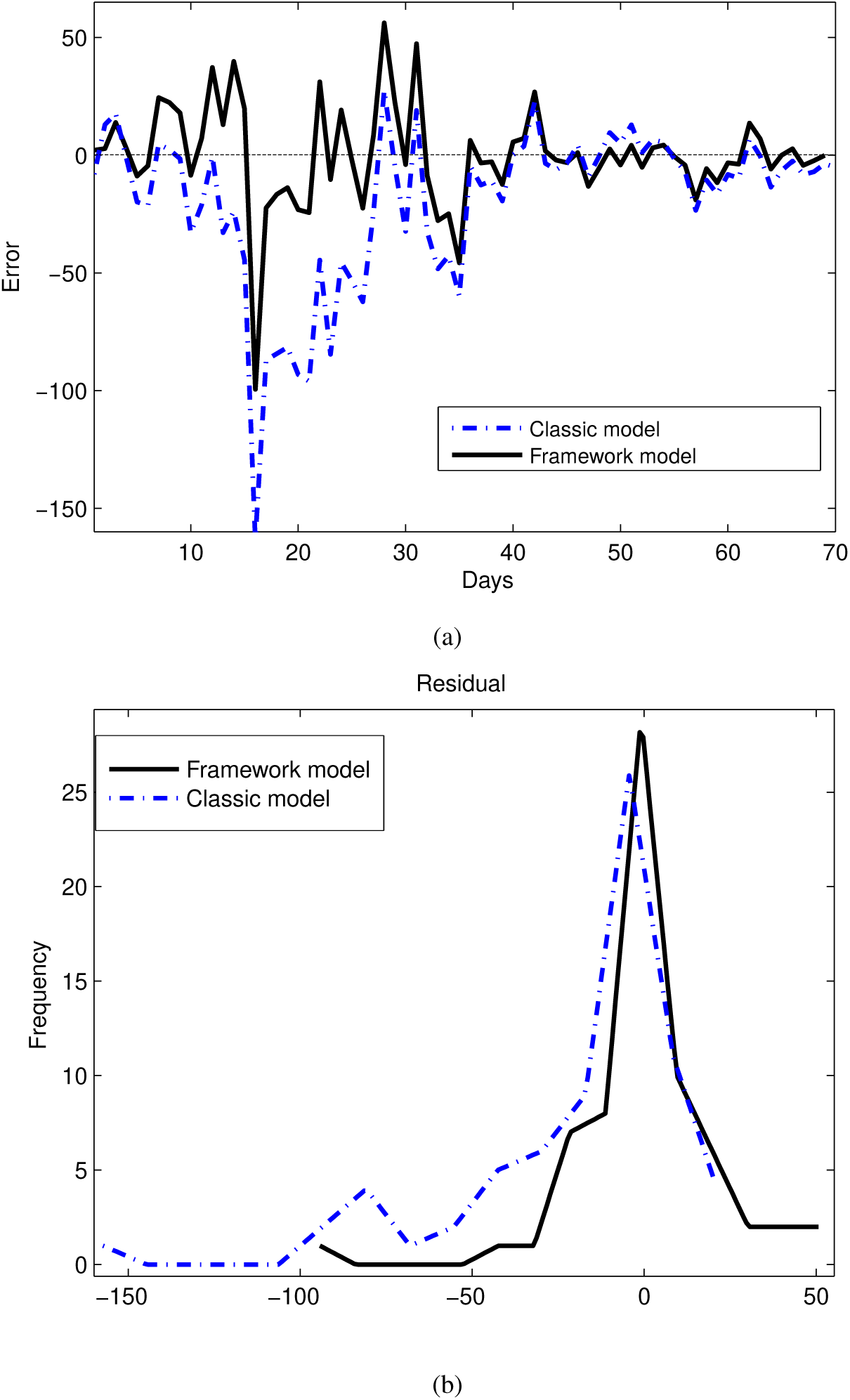
(a) Model residuals. (b) Classical model residuals.

Table (5) quantifies the main statistical measures that characterize the error’s distribution for each model (classic model and our framework), where 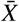 is the average, *M* is the fashion, *std* is the standard deviations and *S* is the distribution bias. The *S* bias is calculated through the second *Pearson coefficient* of the form 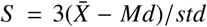, where *Md* It is the median. The coefficient of bias gives an idea of the symmetry of the distribution, if *S* > 0 the symmetry of the distribution is positive, if *S* < 0 the symmetry is negative and if *S* = 0 the distribution is symmetric. This means that the closer to 0 the value of *S* is, the better the approximation of the model is. In both cases, both the model with social behavior (with FCM) and the model without social behavior (without FCM) the value of the bias is very close to 0, with a slight negative asymmetry for the second case. In both the mean and the median, it can be seen that 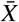 and *M* are considerably lower for the two models based on individuals.

**Table 5:** Statistics of the error

| Modelo | $\bar{X}$ | $M$ | $std$ | $S$ | Kurtosis |
| --- | --- | --- | --- | --- | --- |
| *SEIR* | -19.57 | -9.15 | 54.75 | 0 | 1.78 |
| Model | -21.63 | 1.79 | 46.41 | 0 | 1.75 |

### 5.2. Numerical validation

The model in the proposed framework was numerically validated using the Akaike information criterion (*AIC*). This criterion provides a measure of the quality of the model taking into account the precision and complexity of it. Models that have a *AIC* coefficient within the [1 – 2] range consistently support the structural variation in the data, if the value is within the range [3 – 7] they support the variation considerably structure in the data and if the *AIC* > 10 coefficient does not explain some relevant structural variations in the data. The index is calculated as described in the previous section according to:

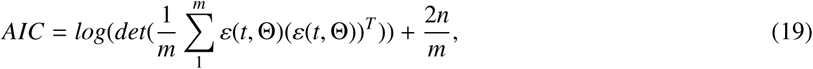

where Θ is the set of *n* uncertainty parameters, *m* is the number of simulations or samples and *ε*(*t*, Θ) the error mesure. The resulting coefficients were *AIC* = 6.1 for the proposed model and for the classic model were *AIC* = 7.5.

Akaike Information Criterion is equivalent to a cross-validation *k-leave-one-out* in longitudinal data models, for example, bandwidth selection (Fang, 2011). This technique is used to evaluate the results of statistical analysis and ensure that they are independent of the partition between training data and the test test. It consists of repeating and calculating the arithmetic average obtained from the evaluation measures on different partitions. It is used in environments where the main objective is prediction and we want to estimate how accurate an implemented model is.

Keeping this in mind we evaluated the statistical significance of these results by calculating the probability that the model’s error better than the error of the classical model. To perform this test we assume the statistical independence of the fitting errors for different data sets and we approach the errors of a *binomial distribution* by a Gaussian distribution. This is possible because we have a high enough number of records for each experiment (5000 data set experiments).The data set was divided into three subsets. First, the proposed and teh classical models were trained with the same data set but with *n* = 10 randomly less data points selected following a uniform probability distribution. With the resulting data set each model was trained in order to obtain 7 generations of parameters. Then, 5000 runs were made for each generation of parameters obtained in the previous step in order to obtain a good approximation of the mean output of the model. Finally, the average error is calculated having in mind the average response of the model using the data that was extracted from the original data set. This process was then carried out for *n* = 20 and *n* = 30. For each set of parameters with different gaps in the data set we test hypothesis *P*(*Error_model_* < *Error_SEIR_*) > *p*.

Table 6 summarizes the results of the statistical significance test for the different validation sets for the model. It can be seen that the implemented model is better than the classical model if the approach error is observed. The confidence interval of the error, in general, is above 90%.

**Table 6:** Errors siginificance for the proposed model

| 10 datos |  |  |  |  |  |
| --- | --- | --- | --- | --- | --- |
| Model | Error | $\mu$ | $\sigma$ | $\frac{\mu_1-\mu_2}{\sigma}$ | $P(p_1 < p_2)$ |
| Proposed model | 8.69 | 0.913 | 0.0046 | 3.58 | 99.55 |
| *S E I R* | 10.04 | 0.896 | 0.005 |  |  |
| 20 datos |  |  |  |  |  |
| Model | Error | $\mu$ | $\sigma$ | $\frac{\mu_1-\mu_2}{\sigma}$ | $P(p_1 < p_2)$ |
| Proposed model | 5.07 | 0.949 | 0.0036 | 1.85 | 88.84 |
| *S E I R* | 5.75 | 0.943 | 0.0038 |  |  |
| 30 datos |  |  |  |  |  |
| Model | Error | $\mu$ | $\sigma$ | $\frac{\mu_1-\mu_2}{\sigma}$ | $P(p_1 < p_2)$ |
| Proposed model | 4.86 | 0.951 | 0.0035 | 2.81 | 97.23 |
| *S E I R* | 5.88 | 0.941 | 0.0038 |  |  |

## 6. Conclusions

The proposed framework allows a transparent system to be implemented transparently and simply. It was demonstrated that with the proposed framework it is possible to adjust and reproduce a real situation with great precision, surpassing even the classic representations of the same system.

The inclusion of behavioral aspects in individuals through *FCM*, perhaps allows the model to capture the dispersion of data throughout the wave and even at the end of the epidemic wave can be seen that the model manages to capture the epidemic outbreaks. Another interesting aspect of the model itself is that it manages to reproduce in a fairly filed way the rapid increase of infected individuals at the beginning of the epidemic. This aspect is not trivial when it comes to modeling this type of phenomenon.

The *FCM’s* output alters the way in which individuals relate to each other. It is likely that due to this perception of the agents at a local and global level, the rapid dynamics of the first stage of the wave as well as the end of the wave will be captured, when the situation improves, individuals tend to recover their movement patterns natural and that is why new smaller shoots arise, typical in this type of systems.

The statistical results show better results for the proposed model if compared against the classical model. This can be seen in the mean and median error made by the model and also in the waste graph.

The homogeneity of contacts could not be assumed in this approach. When modeling space explicitly, this hypothesis is not natural. The use of flocking to model the social behavior of individuals contributes to effective contacts between susceptible and infectious as a result of group formation when it is assumed that individuals are distributed homogeneously in the environment.

## Data Availability

All the data is available in the sources pointed in the paper

